# The Antimicrobial Resistance Campaigns Reach and Community Engagement Evidences from Tanzania’s One Health Initiatives

**DOI:** 10.64898/2026.10.01.26364341

**Authors:** Rajab Awami, Khanafi Said, Jaynes Kabuhaya, Gloria Swilla, Genoveva Ndunguru, Casandra Korosso, Joshua B. Munema, Milka Silvatory Mutashobya, Samweli Charles, Hamisi Msagama

**Affiliations:** One Health Society, Dar es Salaam, Tanzania; Department of Zoology and Wildlife Conservation, College of Natural and Applied Sciences, University of Dar es Salaam, P.O Box 35064, Dar es Salaam, Tanzania; School of Medicine, KCMC University, Moshi, Tanzania

**Keywords:** WAAW, Community engagement, Tanzania, Antimicrobial resistance, awareness campaigns

## Abstract

**Objectives:** Antimicrobial resistance (AMR) is no longer a future health problem but rather a present public health crisis. For over a decade, national AMR awareness initiatives have been undertaken in Tanzania to implement National Action Plans (NAPs). However, the assessment of whether these initiatives have reached the target population is still limited. This study evaluated physical reach, public perception and community participation in national AMR awareness campaigns in two regions in Tanzania.

**Methodology:** This is a quantitative, community-based, cross-sectional study in Dar es Salaam and Mwanza regions, Tanzania. The multistage technique was used to sample 2,672 questionnaire respondents. Participant characteristics and perceptions of the campaign were summarised using descriptive statistics. Multivariable logistic and firth regression was used to establish predictors of high AMR campaign perception and physical campaign reach.

**Results:** Only 81 (3.03%) respondents reported being reached by AMR awareness physical campaigns. Physical reach was strongly linked with high education and skilled occupation. Respondents in Mwanza were less likely to be reached by campaigns compared to Dar es Salaam. Despite limited physical reach, the overall perception campaigns were positive. The majority of the respondents agreed on the importance of community involvement (91.6%), AMR education should be brought into schools (98.9%), believed the campaign messages were accurate (79.6%) and would like to get involved with future campaigns (91.4%). Higher odds for favourable perception were found to be independently associated with physical campaign reach (AOR=2.77, 95% CI: 1.40–5.49).

**Conclusions:** The Promotion of AMR awareness campaigns in Tanzania seems to be well accepted by the public despite limited reach. Future campaigns need to shift away from one - off activities and towards ongoing and inclusive community engagement, leveraging schools, community leaders, community-based structures and systematically tracking reach and behaviours.

## Background

Antimicrobial resistance (AMR) is no longer a future health problem but rather a present public health crisis ^1^. In 2019, bacterial AMR alone was responsible for an estimated 1.27 million deaths worldwide ^1^. Sub-Saharan Africa bears the heaviest share of this silent pandemic reasons being weak surveillance, poor diagnostic access and poor regulation of antimicrobials across One Health domains ^2,3^. In May 2015, the Global Action Plan on Antimicrobial Resistance (GAP-AMR) was endorsed to respond to the AMR problem. Up to date, 178 countries have developed their own multisectoral National Action Plans (NAPs) to align with GAP-AMR recommendations ^4^. GAP-AMR set five strategic objectives, one of which (objective 1) brought about the establishment of World AMR Awareness Week (WAAW) ^5^. WAAW, held every 18– 24 November, gives stakeholders a fixed annual window to run public education activities across the human, animal, and environmental sectors ^5^. How well WAAW and other campaigns actually reach the intended audience is a separate question.

A systematic review across the WHO African regions found that AMR education has expanded steadily over the past decade. Delivery mode is tied to short annual outreaches like WAAW. However, in between, little is happening at the community level ^6^. Global and African reviews of AMR campaigns revealed short-term knowledge gains, poor measurement of coverage equity and the tendency of low- and middle-income countries not to report data on population-level reach ^7,8^. WHO’s global survey of AMR recommendations includes proper definition of the target population and monitoring and evaluation as to whether it was actually reached or not, rather than stopping at message content alone ^9^. Evaluations of awareness programs in Ghana that found knowledge concentrated among healthcare workers and city residents, while rural and less-educated groups were largely missed ^10^. In Zimbabwe, rural pastoral households were found to be missed out by most awareness programmes ^11^.

Tanzania has various policies to respond to AMR. Its first NAP was released in 2017 and perfectly framed by all One Health Ministries ^12^. The current NAP (2023–2028) set awareness objectives and activities through various approaches, WAAW being one of them ^13^. A notable approach is the official translation of AMR into the Swahili word “Usugu wa Vimelea vya Magonjwa Dhidi ya Dawa (UVIDA)”. Despite decades of NAP implementation of creating awareness through various campaigns, evidence of reach, perception and community engagement status remains limited. Existing Tanzanian studies assess general knowledge, attitudes, and practices related to AMR and antibiotic use ^14,15^.

None of the studies assesses the actual reach or community engagement of a national campaign in relation to the demographic characteristics of audiences. Researching on awareness equity matters because reach is normally not a random outcome but rather it reflects a population which has a more social advantage than others. In Tanzania two questions remain unanswered. One, what share of the public has actually been physically reached by national AMR campaigns? Two, how reach, along with community engagement, differs across demographic and geographic groups. Without these evidence, upcoming campaigns will always stay a guess. This study evaluated the reach, public perception and community engagement of national AMR awareness campaigns among residents of Dar es Salaam and Mwanza, Tanzania.

## Methodology

### Study design

This study employed a quantitative, observational, and community-based cross-sectional study design. The cross-sectional design was selected to simultaneously capture the AMR campaign exposure, public perception and their demographic predictors across two regions of Tanzania at a single point in time.

### Study settings and target population

The study was conducted in two administrative regions of mainland Tanzania (Dar es Salaam and Mwanza). Dar es Salaam is a primary commercial metropolis of Tanzania located in the eastern part, which is characterised by high population density, urban and modern infrastructure and concentrated media exposure. Mwanza region is in the lake zones of Tanzania, located in the northwestern part of Tanzania, having a mix of urban, peri-urban and agropastoral communities. The target population of this study was residents aged 18 years and older who resided in the two regions during the survey period.

### Sample size and sampling technique

The sample size aimed to establish adequate precision and compare outcomes across demographic and geographical subgroups. The minimum sample size was calculated using Cochran’s formula for cross-sectional surveys, assuming a 50% response proportion to yield maximum variance, a 95% confidence interval (Z=1.96) and a 2.0% margin of error ^16^. After adjusting for design effects of 1.8 for multi-stage cluster sampling and a 10% non-response rate, a sample size of 2674 respondents was required ^16^.

A multistage sampling technique was used to select participants. Administrative municipal councils in Dar es Salaam and Mwanza were randomly selected. Wards were then chosen using probability proportional to size, and finally, the individual respondents within the public hubs and residential areas were selected using systematic random sampling.

### Inclusion and Exclusion Criteria

Eligible participants were adults aged 18 years or older who had resided in the selected administrative wards for at least six months before the survey. Temporary visitors, individuals unable to provide informed consent, and health professionals directly involved in organising national AMR campaigns were excluded to minimise response bias

### Data Collection procedure

Data were collected using a structured questionnaire that was administered electronically by trained research assistants via Kobo Toolbox. The survey tool was organised into three primary sections (socio-demographic profile, physical AMR campaign reach, and public perception and attitude toward the AMR campaigns).

### Variable definition and operationalisation

The primary dependent variables were physical campaign reach (binary: Yes vs No) and overall AMR campaign perception. The independent predictor variables were categorised as follows: sex (male vs female), age category (youth - 18 - 35 years, adults - 36 -50 years, and seniors > 50 years), education level (no formal education, medium education - primary and secondary education completion and higher education - tertiary/university), occupation (skilled -formal employment, professional and technical vs unskilled - informal sector, small-scale vendors and manual labour).

### Statistical data analysis

Data processing and analysis were conducted using R-Studio version 4.5.0. Categorical variables were summarised as frequencies and percentages, while continuous variables were expressed using means with their corresponding standard deviations or median with their corresponding interquartile ranges.

To measure the overall perception, a summative perception index was created as a composite measure of attitude of respondents to various components of AMR campaigns (campaign delivery, institutional trust, future engagement). The items demonstrated good internal consistency (Cronbach’s α = 0.475) and exploratory factor analysis (KMO = 0.615; Bartlett’s p<0.001) indicated that the items were measuring multiple related domains (campaign delivery, trust, future engagement). The perception scores were then categorised into high perception (score > 5) and low/medium perception (score <5). To evaluate the independent determinants of high campaign perception, a multivariate binary logistic regression model was fitted to yield the adjusted odds ratios and 95% confidence intervals.

For a physical campaign reach, Firth’s penalised likelihood logistic regression was used to analyse its independent predictors. Firth’s model was used because of the smaller proportion of participants who have been reached by physical campaigns. The statistical significance across all models was established at p<0.05

### Ethical considerations

Approval to conduct this survey was given by the Prime Minister’s Office - Regional Administration and Local Government with reference number AH.161/298/03. Before field data collection, additional permission was sought from District Medical, Veterinary and Medical Officers. All of the participants participated voluntarily after signing informed consent. Anonymity and data confidentiality were strictly maintained throughout the data collection and storage. No human samples were collected during the study.

## Results

### General demography of respondents without stratification by region

A total of 2,672 respondents were interviewed, and more than half were male (58.6%). The respondents had a mean age of 32.2±9.7 years. The sample was skewed towards young adults, where three quarters (71.9%) of respondents were under the age of 35 years. There were more participants from Mwanza region (59.7%) compared to Dar es Salaam (DSM) (40.3%). Education levels ranged from uneducated (6.9%) to medium (primary and secondary) (73.1%) and higher education (university and colleges) (20%). The majority of respondents (74.6%) were unskilled working in informal sectors. The demography stratified by region is presented in Table 1.

**Table 1:**
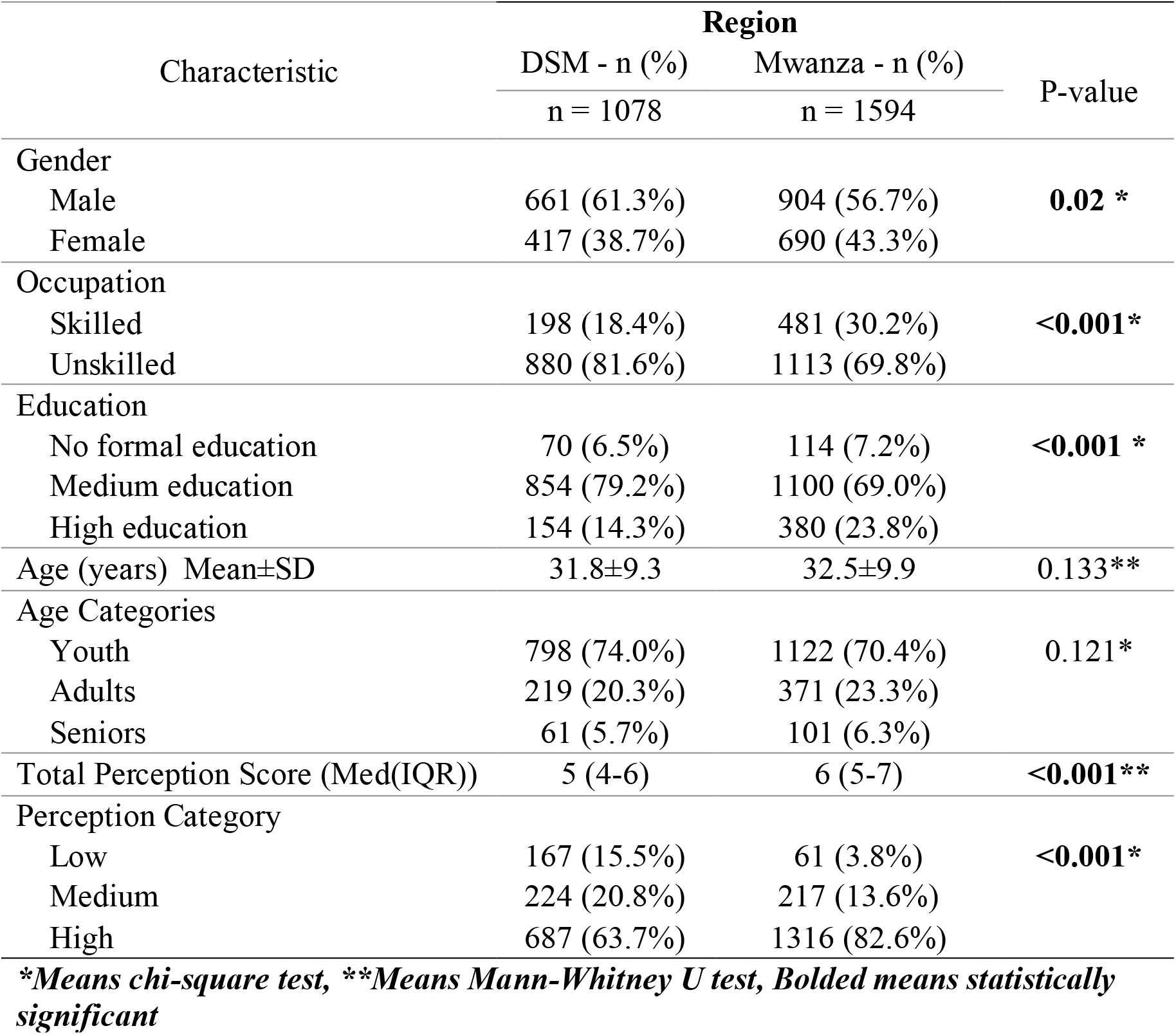
Demographic distribution of respondents stratified by region of residence.

Surprisingly, of all respondents, only 81 respondents (3.03%) reported being reached by a Physical AMR campaign. The majority were male (61.7%, p = 0.637), with a median age of 31.2±9.9 (p=0.211) years. The major portion of those who were reached (75.3%) were youth (18-35 years), while 18.5% were adults (36-50 years), and 6.2% were over seniors of 50 years old, p=0.733. Geographically, a large proportion of reached respondents live in DSM (61.7%) compared to Mwanza (38.3%), p<0.001. Reached participants had a medium education (50.6%), while 48.1% had higher and (1.2%) primary education, p<0.001. There was no observed disparity in AMR physical reach in relation to occupation; the physical campaign reached 50.6% of skilled workers and 49.4% of unskilled workers, p<0.001.

**Table 2:**
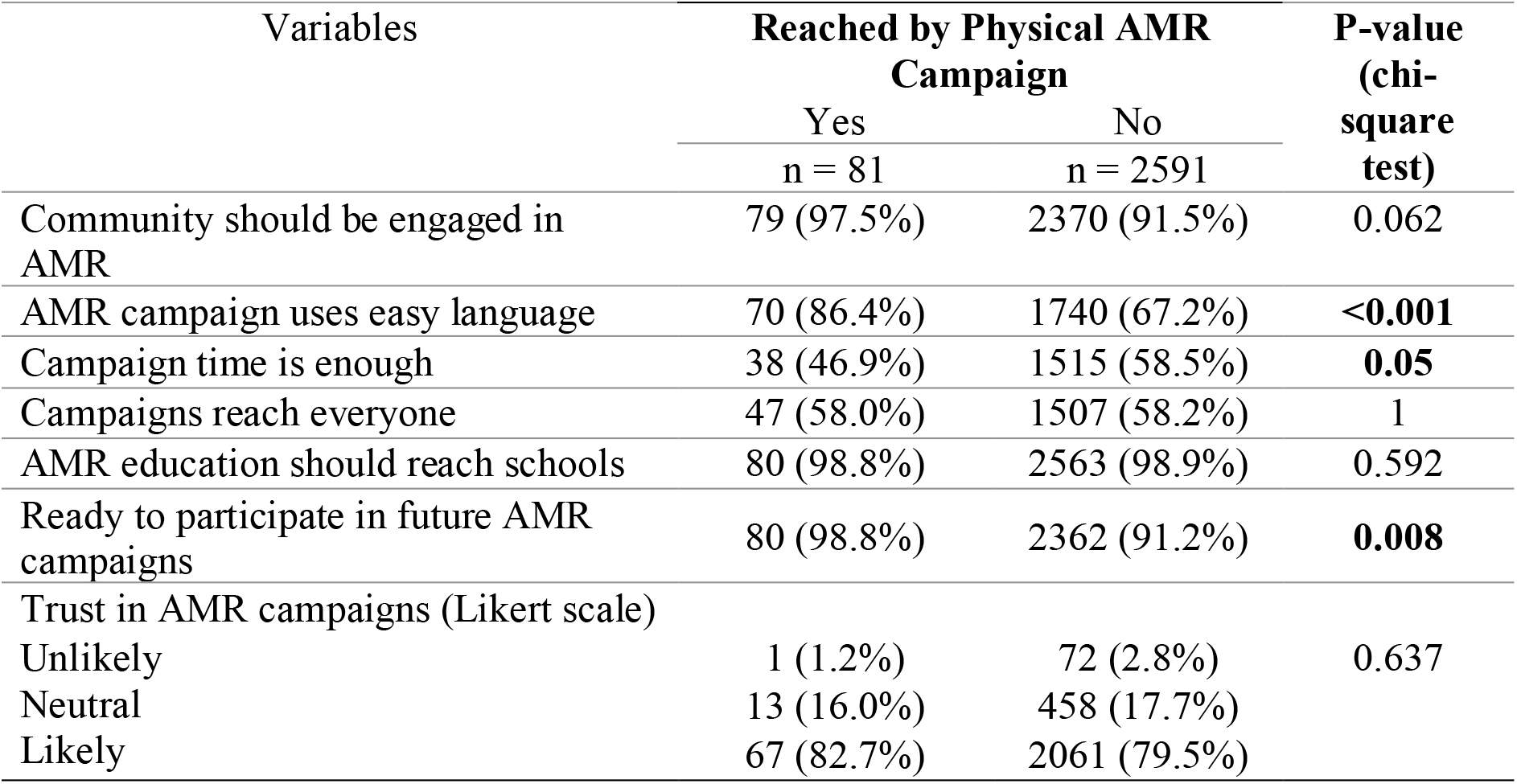
Perception variables stratified by AMR campaign reach status.

Out of 2672 respondents, 2,591 (96.97%) approved not being reached by the AMR campaigns. Males are slightly unreached (1515, 58.5%) than females (1076, 41.5%). The media age was 32.2±9.7 years. Regionally, 1563 (60.3%) resided in Mwanza and 1028 (39.7%) in DSM. The level of education attained was variable, whereas 1913 (73.8%) had medium, 495 (19.1%) higher and 183 (7.1%) no formal education. The majority of the occupations not reached (1859, 71.7%) were youth, while 22.2% were adults and 6.1% were seniors over 50 years old. A large proportion of the unreached were unskilled 1953 (75.4%). The typical unreached group represents youth, male, and Mwanza residents, medium educated and unskilled.

A summative perception score was generated using community engagement, language easiness, campaign reaching everyone, time, trust, school reach and future participation questions. The scores ranged between 1 and 7 (median 6, IQR (4-7), mean 5.46, SD 1.32 and strongly left-skewed -0.5). The scores (n=2672) were categorised as low 1-3 (8.5%), moderate 4 (16.5%), and high 5-7 (75%).

**Figure 1:**
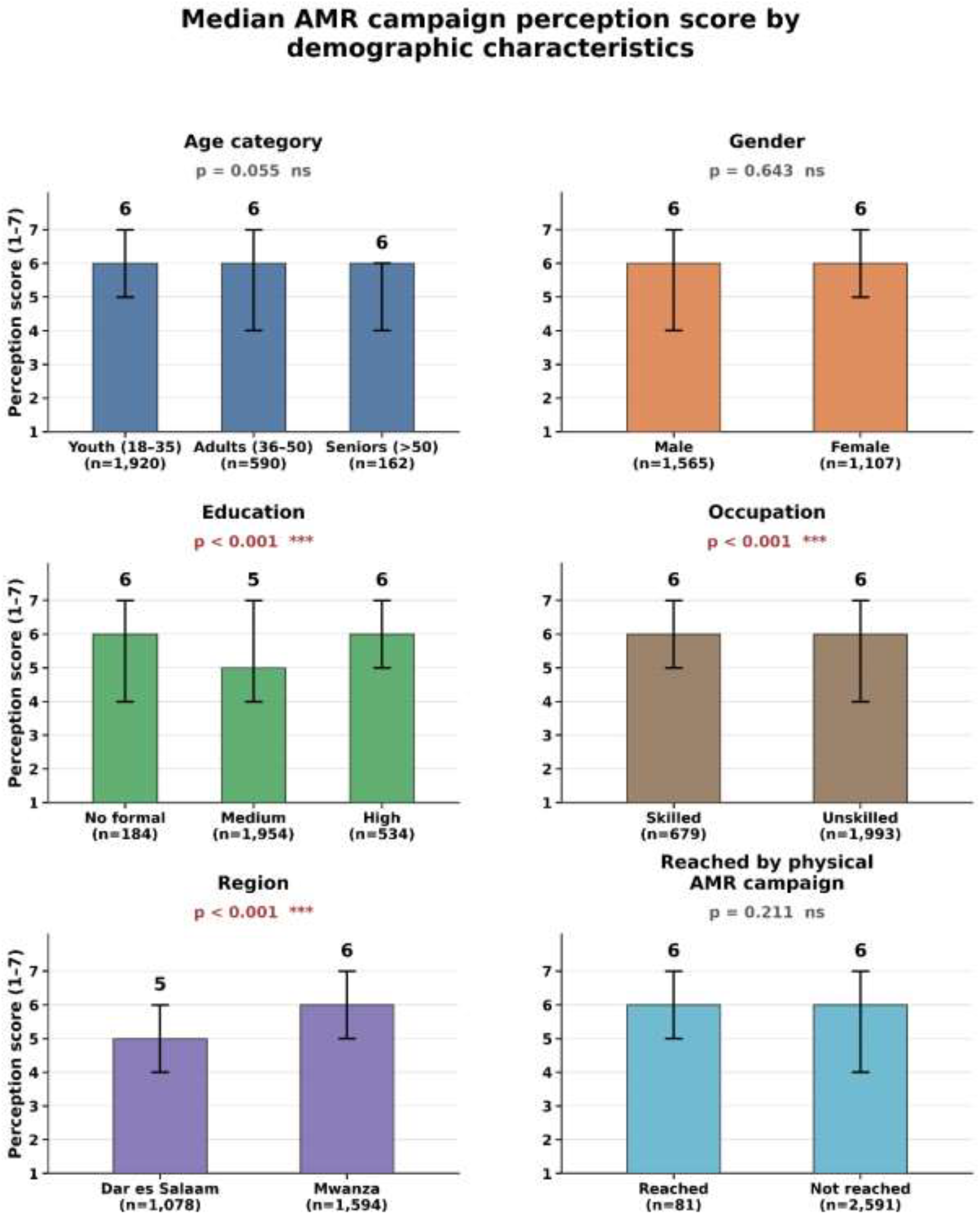
Median perception score faceted with demography (age category, gender, education, occupation, region of residence and being reached by AMR campaign). Scores ranged between 1-7. Intervals show interquartile range (Q1-Q3). P-values generated from Mann-Whitney U test (for 2 groups) or Kruskal-Wallis test (≥3 groups). ns = non-significant and ***p < 0.001.

Respondents responded differently regarding their views on various aspects of WAAW. Almost everyone (91.6%) agreed that communities should be involved in the planning and implementation of AMR campaigns. In terms of campaign communication, 67.7% agreed that the campaign language was easy to understand and more than half (58.1%) showed a positive attitude toward the adequacy of campaign time. The same percentage (58.1%) felt that campaigns are effective at reaching everyone who needs to be educated. Overall, trust in information from AMR campaigns was generally positive, whereas 79.6% were likely to trust AMR campaign messages. Nearly all of the respondents (98.9%) believed that AMR education should reach schools and 91.4% were willing to become part of any future campaign. Regional stratification of these responses is summarised in Table 3.

**Table 4:** Perception variables stratified by region of residence.

| Perception variables | Region |  | P-Value<br>(Chi square test) |
| --- | --- | --- | --- |
|  | Dar es Salaam<br>(n = 1078) | Mwanza<br>(n = 1594) |  |
| Communities should be engaged in AMR campaigns | 1006 (93.3%) | 1443 (90.5%) | **0.013** |
| Campaigns use easy language | 691 (64.1%) | 1119 (70.2%) | **0.001** |
| Campaigns time is enough | 442 (41%) | 1111 (69.7%) | **<0.001** |
| AMR campaigns reach everyone | 470 (43.6%) | 1084 (68%) | **<0.001** |
| I trust AMR campaigns |  |  |  |
| Unlikely | 55 (5.1%) | 18 (1.1%) | **0.001** |
| Neutral | 324 (30.1%) | 147 (9.2%) |  |
| Likely | 699 (64.8%) | 1429 (89.6%) |  |
| AMR education should reach schools | 1070 (99.3%) | 1573 (98.7%) | 0.223 |
| Ready for future participation | 997 (92.5%) | 1445 (90.7%) | 0.112 |

**Figure 2:**
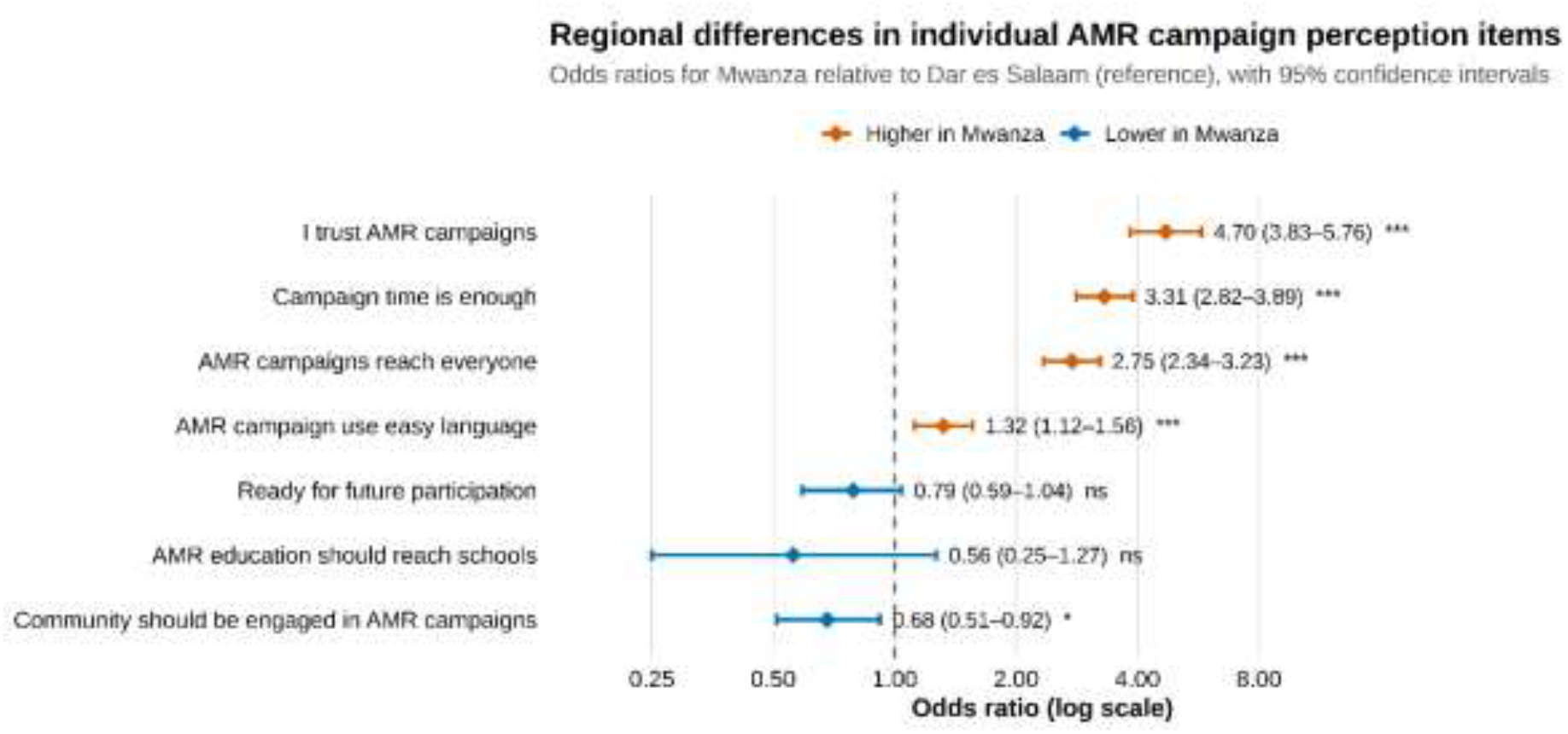
Forest plot of region (Mwanza and DSM (reference)) specific odds ratio (OR) for seven individual AMR campaign perception items. Each OR was estimated from a separate binary logistic regression model (item endorsement as outcome, region as sole predictor; n = 2,672 for all items). Diamonds represent point estimates and horizontal bars represent 95% Wald confidence intervals. The vertical dash line indicates the null value (OR=1). Colour denotes the direction of the regional difference (vermillion = higher odds in Mwanza, blue = lower odds in Mwanza). Significance: *p<0.05, **p<0.01, ***p<0.001, ns = not significant (p>0.05).

The perception variables were tabulated with demography p-values from the chi-square test are presented in table 5.

**Table 5:** Perception variables(Columns) tabulated with demography P-values are from chi-square test.

|  | Respondents Demography |  |  |  |
| --- | --- | --- | --- | --- |
|  | Sex | Occupation | Education | Age |
| Communities should be engaged in AMR campaigns | 0.7 | 0.3 | <0.001 | 0.1 |
| AMR campaigns use easy language | 0.1 | 0.3 | <0.001 | 0.049 |
| Campaign time is enough | 0.9 | <0.001 | <0.001 | 0.54 |
| AMR campaigns reach everyone | 0.3 | 0.003 | 0.69 | 0.2 |
| I trust AMR campaigns | 0.9 | <0.001 | <0.001 | 0.07 |
| AMR education should reach schools | 0.9 | 0.9 | 0.4 | 0.012 |
| Ready for future participation | 0.4 | 0.1 | <0.001 | <0.001 |

**NB: Categories of demography are: Sex (Male & Female), Region of residence (Dar es Salaam & Mwanza), Education categories (higher, medium & low education) and age groups (Youth, Adults and Seniors). Numbers are p-values from the chi-square test.**

Sample segmentation was done by cross tabulating four mutual exclusive groups based on campaign reach and perception categories. The group’s composition includes reached but high perception (n=71, 2.66%), reached but low/medium perception (n=10, 0.37%), not reached but high perception (1,932, 72.31%), and not reached but low/medium perception (n=659, 24.66%). The demographic profile of each segment is summarised in heatmap figure 3.

**Figure 3:**
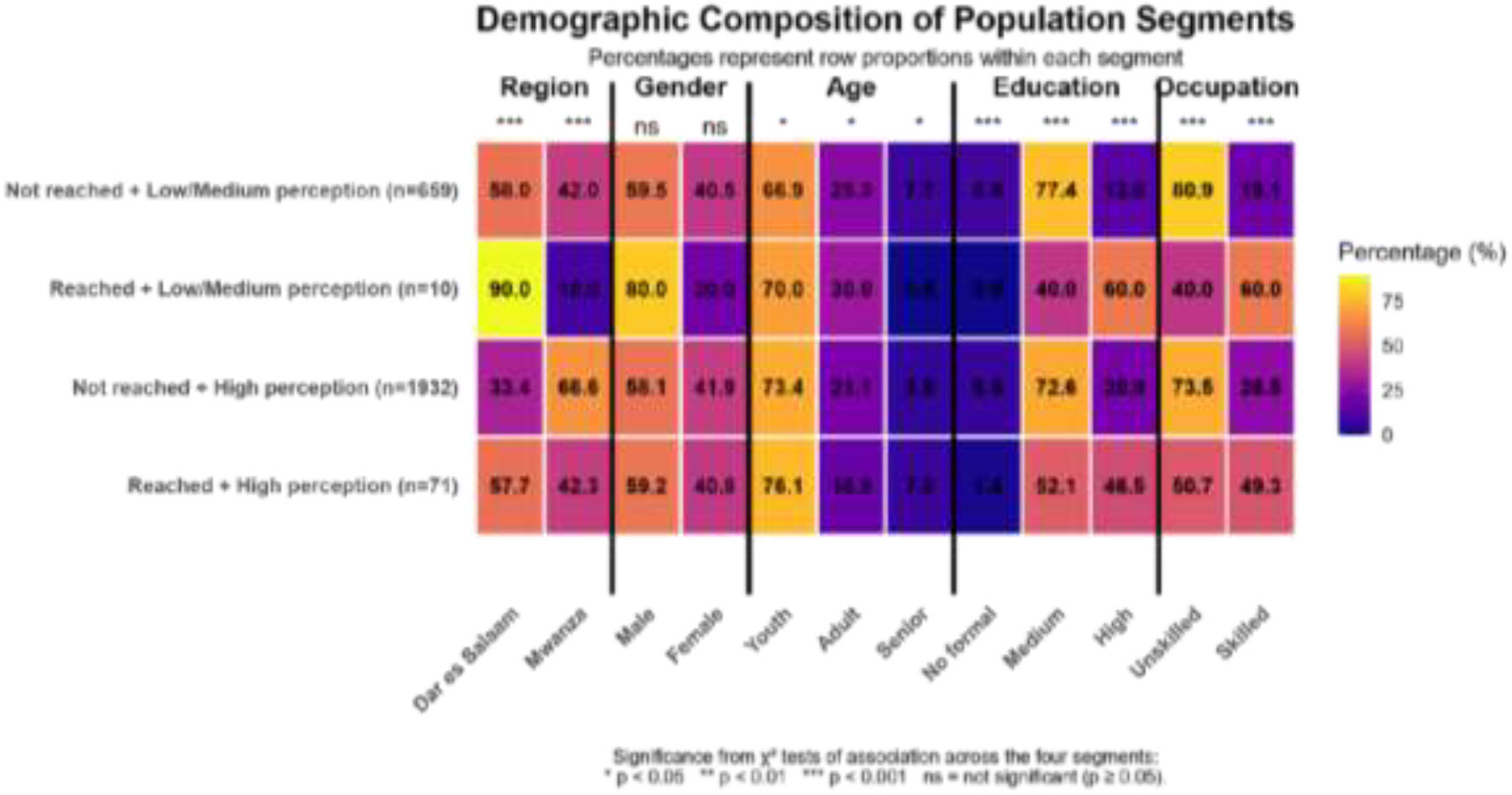
The demographic composition of the population segment presented as a percentage proportion within each segment.

The multivariable logistic regression showed that respondents from Mwanza had significantly higher adjusted odds of high AMR perception (AOR=2.73, 95% CI 2.27–3.28, p<0.001). Respondents who had been physically reached by an AMR had significantly higher adjusted odds of high AMR perception (AOR=2.77, 95% CI 1.40–5.49, p=0.003). After adjusting, adults and seniors were less likely to perceive high risk compared to youth (AOR=0.73, p=0.005 and AOR=0.66, p=0.028, respectively), while gender and education and occupation were not independent predictors. With a pseudo-R^2^ of 0.052, the model had a modest level of explained variance, and was not a victim of multicollinearity (VIF<3.6 for all terms) with a Hosmer–Lemeshow p<0.92. The results of the AOR are presented in figure 4.

**Figure 4:**
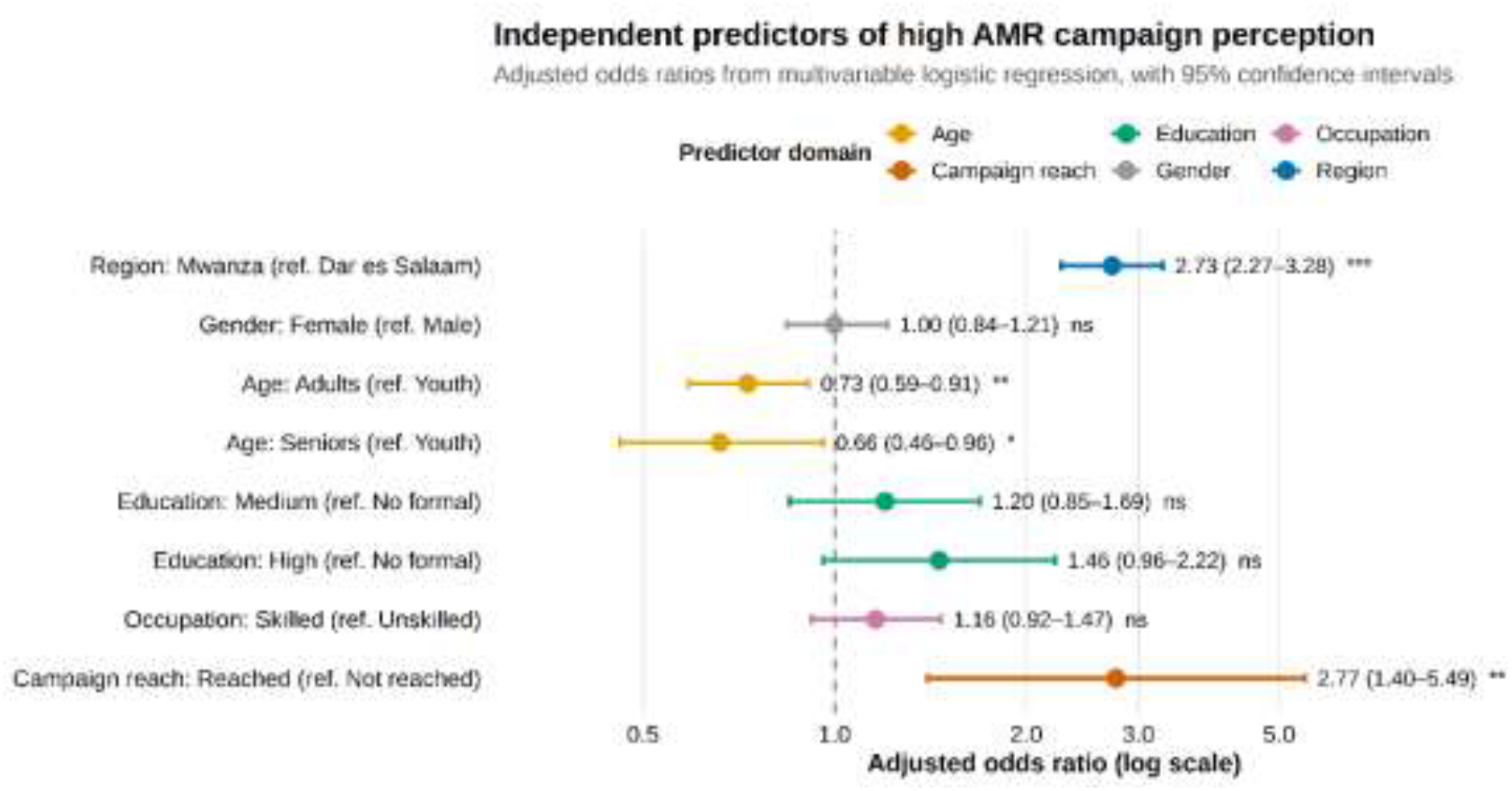
Forest plot of Adjusted Odds Ratios (AOR) for high perception (vs. low/medium perception) from a single multivariate logistic regression model, including region, gender, age category, education category, occupation and campaign reach status (n = 2,672; Hosmer-Lem show p = 0.92; max VIF = 3.57). Diamonds represent AOR and horizontal bars represent 95% wild confidence intervals. The vertical dash line indicates the null value (AOR=1). Colour denotes the predictor domain. Significance: *p<0.05, **p<0.01, ***p<0.001, ns = not significant (p>0.05)

The survey data was sparse (only 81 reached out of 2672) and hence firth’s penalised logistic regression was used. Skilled occupation (AOR=2.37, 95% CI 1.45–3.87, p<0.001) and high education (AOR=7.62, 95% CI 1.44–40.26, p=0.017) were independently associated with greater odds of reach, while Mwanza residents were substantially less likely to have been reached than Dar es Salaam residents (AOR=0.30, 95% CI 0.19–0.47, p<0.001). The results of a logistic regression analysis (standard vs firth) of predictors of AMR campaign reach are summarised in figure 5.

**Figure 5:**
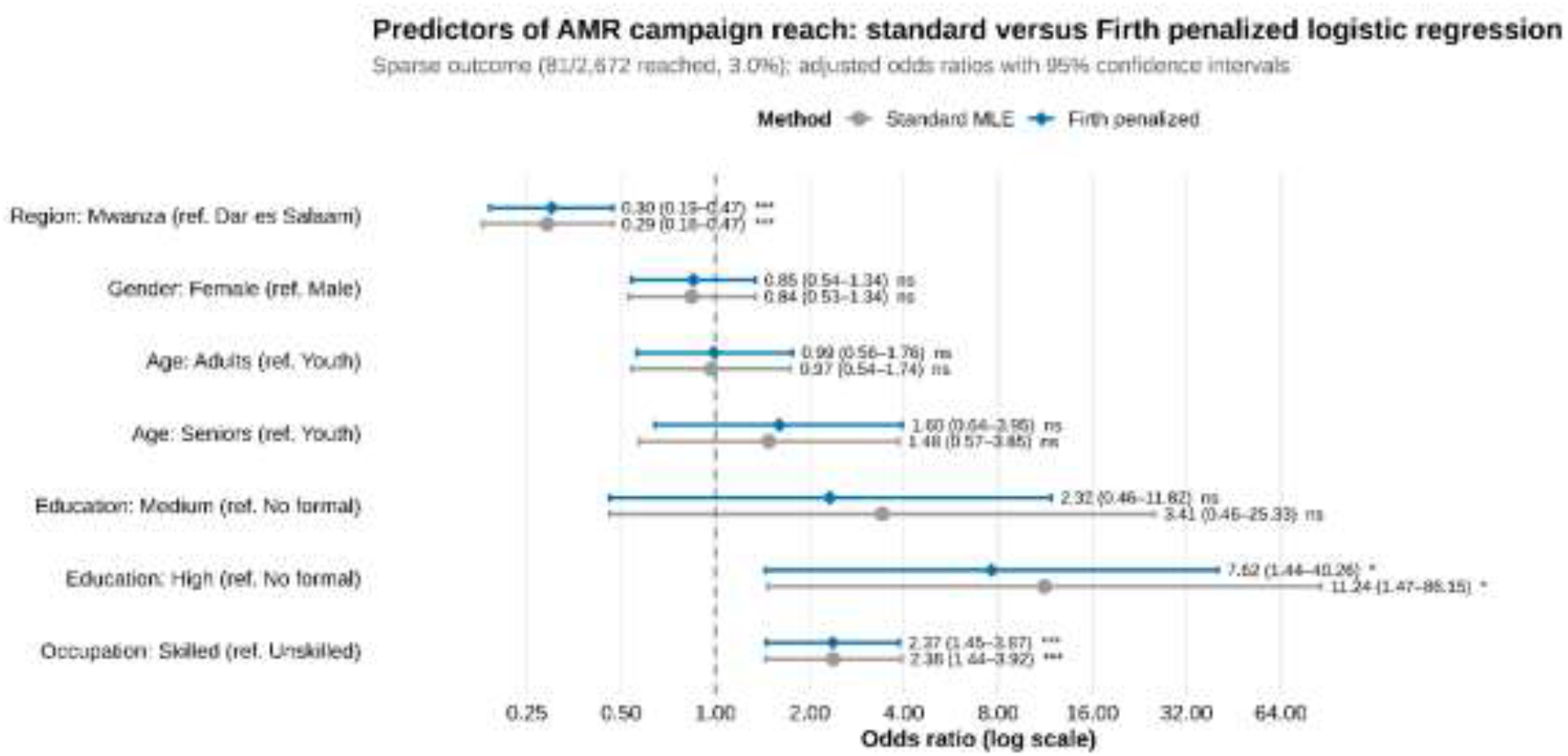
Comparison of Adjusted Odds Ratios (AOR) for predictors of physical AMR campaign reach, which were estimated by using standard maximum-likelihood logistic regression (see grey circles) versus Firth bias reduced penalised logistic regression (blue diamonds), both adjusted for region, gender, age category, education category, and occupation simultaneously (n = 2672, only 81 were reached). The vertical dash line indicates the null value (AOR=1). Significance from the firth model: *p<0.05, **p<0.01, ***p<0.001, ns = not significant (p>0.05).

## Discussion

To the best of our knowledge on the available evidence, this is the first study in Tanzania to assess whether respondents were physically reached by AMR awareness campaigns alongside their attitudes towards the campaigns. The existing studies mainly focused on antimicrobial awareness and its use ^14,17^. Drawing respondents from the top two most populated regions in Tanzania, nearly three quarters of those not physically reached by physical AMR campaigns were youth. This is consistent with Tanzania’s population structure, where more than three quarters are below the age of 35 years ^18^.

Our findings reveal a mismatch, attitudes toward AMR messaging and participation remain highly favourable despite extremely low physical campaign reach. This mismatch is similar to other Tanzanian studies, where there is reported high AMR awareness despite limited structured community campaign reach ^14^. The WHO Africa Region is reported to consistently prioritise targeted community-based interventions and education on paper ^6^. However, inconsistent in implementation and monitoring at delivery points leading to uneven population coverage ^6^. This disconnect between community communication intention and actual delivery reflects a broader recurring gap in AMR reach and messaging across the region.

High perception scores among three quarters of unreached groups show perceptions and knowledge are shaped beyond direct reach alone. A similar situation is explained by the diffusion theory and the independent influence of socioeconomic factors, housing conditions, and general health knowledge among respondents ^19-21^. Campaigns combining mass media with healthcare provider interaction are reported to be consistently effective in strengthening awareness and perception, while direct participation may have minor effects on attitudes ^22,23^.

Interestingly, respondents from Mwanza had higher perception scores than those in Dar es Salaam despite having almost no physical campaign. This suggests that perceptions and attitudes are heavily influenced by indirect activities, such as routine healthcare services and non-formal community-level information circulation, similar to direct participation in formal interventions ^22^. Educated and skilled respondents were more likely to report AMR campaign reach. This is consistent with other studies, education and socioeconomic status strongly predict AMR knowledge and awareness in Tanzania ^14,17^. However, after adjusting for other factors, youth showed more favourable perception scores than senior adults, contrary to studies which associated older age with higher AMR knowledge ^14^. This difference suggests that factors shaping receptiveness to campaigns are not the same as those driving actual AMR knowledge. Almost all respondents are ready to participate in physical campaigns, showing strong support for community and school-based AMR education. This proves that communities are ready for expanded outreach programs and campaigns. However, these should be inclusive, since high-risk groups such as livestock-keeping households are frequently overlooked by most AMR campaigns ^24^. Furthermore, awareness alone is insufficient because Tanzanian health providers’ stewardship practices are heavily limited by systemic and resource constraints ^25,26^. Therefore, future AMR expansion programs must actively combine public awareness with direct investments in regulations, healthcare systems, and One Health strategies.

### Strength and Limitations

This is one of the first studies in Tanzania to conduct an evaluation of AMR awareness campaigns by examining the campaign feedback loop and its potential to improve public knowledge of AMR. The study provides valuable evidence of participants’ attitudes and behaviours following exposure to AMR awareness campaigns. The findings contribute to the limited body of evidence on the effectiveness of AMR public awareness interventions in Tanzania and beyond. It can inform the design of future campaigns and policy decisions.

Despite its strengths, the study has several limitations. Data were collected from only two regions of Tanzania. Given that the country comprises 31 regions, including five in Zanzibar, the findings may not be fully representative of the national population, limiting their generalisability. Also, the study population was disproportionately composed of young adults under the age of 35 years. As a result, the perspectives and experiences of older adults may be underrepresented, which may affect the applicability of the findings across all age groups.

## Conclusion

This study revealed a significant difference between the acceptance of AMR awareness campaigns and the actual reach in Dar es Salaam and Mwanza, Tanzania. Only 3.03% were physically engaged despite the overall perception and desire being was high. Among the respondents, a high level of education, being Dar es Salaam residents and skilled occupations, had a higher physical reach. The results indicate key inequities in how AMR awareness activities are currently being delivered. Future campaigns need to shift away from one-off activities and towards ongoing and inclusive community engagement, leveraging schools, community leaders, community-based structures and systematically tracking reach and behaviours. The evaluation of a campaign needs to look at population reach and equity, understanding and behaviour changes to ensure that the awareness-raising effort has led to real and lasting community engagement.

## Data Availability

All data produced in the present study are available upon reasonable request to the authors

## Recommendations

In accordance with our findings below are the actions we propose to improve the reach and impact of future AMR awareness campaigns in Tanzania.

- This study recommends that, instead of having event-based awareness campaigns like the WAAW, there is a need to move to sustainable and continuous community awareness programs that will be integrated into year-round outreach programs. This is important because it will help to maintain public awareness and provide consistent AMR messaging.
- Future interventions should explicitly design communication materials and delivery channels that are accessible to those in rural areas, lower levels of formal education and workers in the informal sector. This is because relying on the general media favours urban, skilled and highly educated populations.
- Since the study noted the high level and willingness of the community to participate in AMR awareness campaigns, more efforts should be made to partner with local leaders, community health workers and schools to disseminate AMR education.
- There should be the adoption of evaluation frameworks capable of tracking behaviour changes across diverse demographic groups to track whether the received information is understood and acted upon.

## Acknowledgement

We are so grateful to all respondents in DSM and Mwanza regions who gave their time to participate in this study. We also thank the field data collectors and research assistants for their hard work during data collection. We appreciate DSM and Mwanza regional and district authorities for their collaboration and for granting permission to conduct this research in their areas of jurisdiction. We also acknowledge the Food and Agriculture Organization (FAO) for their financial support.

## Author’s contribution

**RA:** Study Conceptualization, Investigation, Formal Analysis, Data Presentation, Supervision, Manuscript Review Writing the Original Draft, Acknowledgements, Manuscript Review and Editing, Final Manuscript Checks and Submission. **KS:** Study Conceptualization, Methodology, Formal Analysis, Writing, Manuscript Review and Editing. **JBM:** Writing the Original Draft, Manuscript Review and Editing. **GN:** Writing the Original Draft, Manuscript Review and Editing. **CK:** Study Conceptualization, Writing the Original Draft and Manuscript Review and Editing. **JK:** Study Conceptualization, Writing the Original Draft and Manuscript Review and Editing. **GES:** Study Conceptualization, Writing the Original Draft, Manuscript Review and Editing. **MM:** Writing the Original Draft, Acknowledgements, Manuscript Review and Editing. **SC:** Data Collection, Supervision, Writing the Original Draft, Acknowledgements, Manuscript Review and Editing. **HM:** Data Collection, Supervision, Writing the Original Draft, Acknowledgements, Manuscript Review and Editing. All authors reviewed, revised and approved the final manuscript before submission.

## References

1. Murray CJ, Ikuta KS, Sharara F, et al. Global burden of bacterial antimicrobial resistance in 2019: a systematic analysis. Elsevier B.V.; 2022. p. 629–655.

2. Totaro V, Guido G, Cotugno S, et al. Antimicrobial Resistance in Sub-Saharan Africa: A Comprehensive Landscape Review. The American Journal of Tropical Medicine and Hygiene. 06 Aug. 2025 2025;113(2):253–263. doi:10.4269/ajtmh.25-0035

3. Bennett C, Russel W, Upton R, Frey F, Taye B. Social and ecological determinants of antimicrobial resistance in Africa: a systematic review of epidemiological evidence. Antimicrobial Stewardship & Healthcare Epidemiology. 2024;4(1):e119. e119. doi:10.1017/ash.2024.375

4. WHO. Library of AMR national action plans. WHO. Accessed 20 February, 2026. https://www.who.int/teams/surveillance-prevention-control-AMR/national-action-plan-monitoring-evaluation/library-of-national-action-plans

5. WHO. Global Action Plan on Antimicrobial Resistance. Accessed 20 August, 2026. https://www.who.int/publications/i/item/9789241509763

6. Fuller W, Kapona O, Aboderin AO, et al. Education and awareness on antimicrobial resistance in the WHO African region: a systematic review. Antibiotics. 2023;12(11):1613–1613.

7. Gilham EL, Pearce-Smith N, Carter V, Ashiru-Oredope D. Assessment of global antimicrobial resistance campaigns conducted to improve public awareness and antimicrobial use behaviours: a rapid systematic review. BMC Public Health. 2024/02/06 2024;24(1):396. doi:10.1186/s12889-024-17766-w

8. Maduko W, Olamijuwon E, Kesby M, Hale JM. Public-targeted interventions addressing antimicrobial resistance and antibiotic use in Sub-Saharan Africa: a scoping review. BMJ Global Health. 2025;10(3):e017455. doi:10.1136/bmjgh-2024-017455

9. Huttner B, Saam M, Moja L, et al. How to improve antibiotic awareness campaigns: findings of a WHO global survey. BMJ Global Health. 2019;4(3):e001239. doi:10.1136/bmjgh-2018-001239

10. Loglo AD, Nkrumah-Appau M, Deke J, et al. Exploring and enhancing community awareness on antimicrobial resistance: perspectives from world antimicrobial resistance awareness week celebration in Ghana. BMC Public Health. 2026/01/27 2026;26(1):673. doi:10.1186/s12889-026-26385-6

11. Kembo G, Macheka L, Dembedza MP, Mugadza DT, Manditsera FA. Antimicrobial resistance awareness and food safety concerns among rural households in Zimbabwe. One Health. 2025/12/01/2025;21:101208. doi:10.1016/j.onehlt.2025.101208

12. National Action Plan on Antimicrobial Resistance (2017 - 2022) (Tanzania government) (2017).

13. National Action Plan on Antimicrobial Resistance (2023 - 2028) (Tanzania government) (2022).

14. Sindato C, Mboera LEG, Katale BZ, et al. Knowledge, attitudes and practices regarding antimicrobial use and resistance among communities of Ilala, Kilosa and Kibaha districts of Tanzania. Antimicrobial Resistance & Infection Control. 2020/12/07 2020;9(1):194. doi:10.1186/s13756-020-00862-y

15. Mramba RP, Mbinda AG, Massawe JI. Assessment of public awareness on antimicrobial resistance and practices for antimicrobial use in Dodoma region of Tanzania. Discover Social Science and Health. 2025/02/25 2025;5(1):25. doi:10.1007/s44155-025-00171-y

16. Cochran WG. Sampling Technique. John Wiley & Sons; 1977.

17. Mramba R, Mbinda AG, Massawe J. Assessment of public awareness on antimicrobial resistance and practices for antimicrobial use in Dodoma region of Tanzania. Discover Social Science and Health. 2025-02-25 2025;5 doi:10.1007/s44155-025-00171-y

18. NBS. The report on Population Census in Tanzania Constituencies. United Republic of Tanzania. Accessed 03 March, 2026. https://sensa.nbs.go.tz/publication/majimbo3a.pdf

19. Shutt A, Ashiru-Oredope D, Price J, et al. The intersection of the social determinants of health and antimicrobial resistance in human populations: a systematic review. BMJ Global Health. 2025-05-01 2025;10 doi:10.1136/bmjgh-2024-017389

20. Shutt A, Mukherjee A, Nampoothiri V, Holmes A, Charani E. Social determinants of health and knowledge, attitude and practice studies on antimicrobial resistance—an evidence review and future direction. JAC-Antimicrobial Resistance. 2025-09-04 2025;7 doi:10.1093/jacamr/dlaf141

21. Shutt AE, Ashiru-Oredope D, Price J, et al. The intersection of the social determinants of health and antimicrobial resistance in human populations: a systematic review. BMJ Global Health. 2025;10(5):e017389. doi:10.1136/bmjgh-2024-017389

22. Haenssgen M, Xayavong T, Charoenboon N, Warapikuptanun P, Zaw YK. The Consequences of AMR Education and Awareness Raising: Outputs, Outcomes, and Behavioural Impacts of an Antibiotic-Related Educational Activity in Lao PDR. Antibiotics. 2018-09-03 2018;7 doi:10.3390/antibiotics7040095

23. Gilham E, Pearce-Smith N, Carter V, Ashiru-Oredope D. Assessment of global antimicrobial resistance campaigns conducted to improve public awareness and antimicrobial use behaviours: a rapid systematic review. BMC Public Health. 2024-02-06 2024;24 doi:10.1186/s12889-024-17766-w

24. Kembo G, Macheka L, Dembedza M, Mugadza D, Manditsera F. Antimicrobial resistance awareness and food safety concerns among rural households in Zimbabwe. One Health. 2025-09-01 2025;21 doi:10.1016/j.onehlt.2025.101208

25. Ndaki P, Mwanga J, Mushi M, et al. Practices and motives behind antibiotics provision in drug outlets in Tanzania: A qualitative study. PLOS ONE. 2023-08-31 2023;18 doi:10.1371/journal.pone.0290638

26. Virhia J, Gilmour M, Russell C, et al. “If You Do Not Take the Medicine and Complete the Dose…It Could Cause You More Trouble”: Bringing Awareness, Local Knowledge and Experience into Antimicrobial Stewardship in Tanzania. Antibiotics. 2023-01-25 2023;12 doi:10.3390/antibiotics12020243

